# Effects of HIV prevention interventions on the behaviors of students at the university campuses of Yaoundé (Cameroon)

**DOI:** 10.1101/2025.05.17.25327831

**Authors:** Mohamadou Adama, Linda Esso, Tatiana Mossus, Fabrice Zobel Lekeumo Cheuyem, Hamida Yacoubou, Edwige Omona Guissana, Serges C. Billong

**Author notes:** Corresponding author’s address: Mohamadou Adama, MD, MPH, Department of Public Health, Faculty of Medicine & Biomedical Sciences, The University of Yaoundé I, PO Box 1364 Yaoundé, Cameroon.

## Abstract

**Background:** HIV infection persists at very high levels compared to other parts of the world. It devastates youth and thus jeopardizes the development of countries. This indicates that the success of numerous interventions aimed at changing the sexual behaviors of adolescents and young people could be questioned. These interventions often focus on the individual, and less on the social and cultural environments in which young people’s understanding of sexuality is formed.

**Objectives:** The objectives of this study were to obtain updated baseline data allowing for a better understanding of the HIV epidemic and its determinants and prevention, as well as the evolution of student behaviors following implemented interventions.

**Methods:** To this end, a descriptive, cross-sectional study with an evaluative aim was conducted among students on the campuses of the University of Yaoundé I, the Catholic University of Central Africa, and the Siantou Higher Institute, from May to July 2024. Inclusion criteria were: students aged 18 to 30 years inclusive, regularly enrolled in the aforementioned universities and institutes. We used consecutive recruitment and obtained a total sample size of 1200 students. The collected data were entered and analyzed using IBM SPSS version 26 software. A p-value < 0.05 was considered statistically significant, and confidence intervals were estimated at a 95% confidence level.

**Results:** A total of 1200 students were surveyed during this study, including 572 men and 628 women. The age group under 20 years represented 49.0% of the sample, 21-24 years 40%, and 25 years and older 11%. Regarding the level of education, 98% of students were enrolled in undergraduate programs, and 2% were in postgraduate programs. The majority of students, 99%, resided off-campus, with 75% living with family and 26% living outside the family home with friends or in student housing. Concerning socio-financial characteristics, 92% of students had their parents as their main material/financial support. Among these students, 65% found this material support to be regular, and 48% found it insufficient. Finally, 26% of students reported having a part-time job to support their educational needs. Slightly more than half of the students, 69%, reported having consumed alcohol in the past 12 months, with 27% consuming alcohol several times a month. A little over half, 57%, reported never having consumed alcohol, and 18% consumed alcohol once a month. The majority of students, 85%, reported respectively never having consumed alcohol in a way that prevented them from fulfilling academic obligations or being unable to remember what happened.

**Conclusion:** It appears necessary to redefine the objectives and reformulate prevention programs by strengthening the implemented strategies that have proven effective; however, this should be done with an emphasis on condom use.

## Introduction

Globally, infection with the Human Immunodeficiency Virus (HIV) is no longer among the leading causes of death [1], but it remains the leading cause of mortality among young people and adults in Africa [2]. This situation makes it a region of high endemicity. In West and Central Africa (WCA), Cameroon is the second country after Nigeria bearing the heaviest burden of the HIV epidemic [2]. Despite the significance of the pandemic, the country has adhered to the strategic vision of the Joint United Nations Programme on HIV/AIDS (UNAIDS), which aims to achieve the 95/95/95 targets by 2030 [2]. Cameroon’s national strategy for combating AIDS is based on reducing new infections, particularly among adolescents and young people, through a multisectoral and decentralized approach [3].

The evolution of the HIV epidemic in Cameroon is variable. A thorough analysis of the various national strategic plans developed and implemented has made it possible to describe the evolution of decision-making paradigms in the fight against HIV and AIDS, to highlight the progress made, and to identify the challenges to be addressed for better results [1]. This analysis revealed an improvement in the involvement of sectors other than health and an effective decentralization of interventions over time. Although HIV prevalence has declined, it has remained high in certain risk groups, but the contribution of universities to new infections is suspected to be very high [1, 2]. According to a study conducted by Billong et al., the HIV prevalence in the university population of Yaoundé was 0.8% in 2017 [3]. A sectoral plan to combat HIV was subsequently implemented, targeting students and teachers.

Behavior changes and HIV incidence reduction interventions have been carried out by the National AIDS Control Committee (CNLS) in partnership with the Ministry of Higher Education (MINESUP) since 2017, but little information is available on the HIV epidemic in the Cameroonian university environment. Following these interventions, MINESUP identified priorities during the development of the MINESUP sectoral health strategy for the period 2020-2023, which were articulated into four impact outcomes, namely [4]: by 2023, new infections are reduced by 60% in the university environment and in the central services of the Ministry of Higher Education; by 2023, mortality and morbidity related to HIV are reduced by 70% in the University environment; by 2023, the quality of life of people infected and/or affected by HIV in the university community is improved by 50%; by 2023, the quality of governance of the university response has progressed by 50%.

Five years after the first evaluation conducted by Billong *et al*. on the “HIV Prevalence among Students of the Universities of Yaoundé and Associated Behaviors” [2], and after the interventions carried out by the Ministry of Higher Education targeting students on the university campuses of Yaoundé, it seemed important to us to evaluate the current behaviors of students on the University Campuses of Yaoundé based on socio-economic characteristics and to identify the factors likely to influence them.

## Methodology

### Study design and period

This cross-sectional descriptive study with evaluative purposes was conducted from May to July 2024.

### Study setting

The study was conducted in the Centre Region of Cameroon, specifically on the campuses of the University of Yaoundé I, the Catholic University of Central Africa, and the Siantou University Institute.

### Study population

The study population comprised students enrolled at the aforementioned universities in Yaoundé who agreed to undergo HIV testing.

### Sampling method and sample size

The sites were selected based on purposive sampling, while participant recruitment employed a consecutive sampling method. As the study was cross-sectional with an evaluative aim, the formula used to calculate the minimum sample size was the following COCHRAN formula [32]: N = *Z*2 *X P X QD*2, where: N is the minimum sample size for the study; Z is the Z score at a 95% confidence interval (1.96); P is the prevalence of HIV in the university population, which was 0.8% in 2017; Q corresponds to the value 1 – P (0.992); D is the precision of the study with a 17.5% margin of error (0.175). N = 963 students. Considering documented experiences in similar contexts, we aimed to recruit a larger sample to account for potential dropouts and improve statistical power.

### Inclusion Criteria

Students aged 18 to 45 years, enrolled at the University of Yaoundé I, the Catholic University of Central Africa, or the Siantou University Institute, who provided their informed consent to participate were included.

### Data collection tools and procedure

Data were collected using a pre-tested, interviewer-administered questionnaire that included both closed and open-ended questions. The questionnaire was structured into four main sections, one of which employed a grid format to assess behaviors, attitudes, and practices. Additionally, the Alcohol Use Disorders Identification Test (AUDIT) questionnaire was administered to evaluate alcohol consumption patterns. Data collection was conducted by the principal investigator on the selected university campuses. Prior to data collection, students were informed and mobilized through faculty forums via a press release detailing the study’s purpose, eligibility criteria, and ethical principles. Interviews were conducted in private, designated spaces provided by the university administrations. Participants were seated comfortably, informed about the study, and then administered the questionnaire. Responses were recorded directly onto the questionnaire. Participants were then directed to undergo HIV testing according to the national algorithm, including pre-test and post-test counselling. The questionnaire was anonymized to ensure confidentiality and facilitate follow-up care if necessary. The questionnaire, designed to take approximately 20 minutes to complete, was pre-tested to ensure clarity and validity. A documentary review was conducted to analyze the results of the second objective of our study in document for National Aids Control Committee (NACC). Therefore, to enable MINESUP to Stepping up HIV Response Interventions Led by MINESUP (Ministry of Higher Education) during the period 2018-2023 proved necessary: (1) behavior change communication (BCC)/ Communication for development (C4D) through the training of trainers in interpersonal and mass communication in the university setting; (2) awareness campaigns during special events of commemorative days and national holidays, university games, University Festival of Art and Culture (UNIFAC); (3) capacity building for stakeholders in the university community on HIV, AIDS, and Sexually transmitted infection (STIs), Sexual and reproductive health(SRH), Prevention of mother-to-child transmission(PMTCT), Gender-based violence (GBV), hepatitis, comprehensive care of Person living with HIV (PLHIV); (4) Establishment of health services adapted to adolescents and youth within the health centers of universities and the infirmaries of other Higher Education institutions; (5) Advocacy for concerted action against Gender-based violence (GBV); (6) Information, Education, and Communication (IEC) and capacity building for providers in university health centers and infirmaries on Accidental exposure to blood (AEB) and sexual violence; (7) Establishment of a referral and counter-referral system to specialist doctors and health facilities for the management of co-infections and opportunistic infections, as well as cases requiring hospitalization; (8) Establishment of support groups and networks of PLHIV within universities, private higher education institutions (IPES), university institutions with special status, and central services by psychosocial support workers from the health centers; (9) Integration of the health and HIV component into the strategic planning of the Ministry, Universities, institutions with special status, and IPES; (10) Mobilization of resources related to health and the fight against HIV, AIDS, and STIs; (11) Harmonization of the strategic information system; (12) Promotion and funding of research on HIV, AIDS, and STIs; (12) Interventions will target stakeholders according to their levels of influence.in order to evaluate their effects or impacts.

### Statistical analyses

Data processing and cleaning were performed. The collected data were entered and analyzed using SPSS Version 22. The data analysis was carried out following 2 processes: firstly, we carried out a descriptive analysis of the results and then secondly a bivariate analysis using the chi-square test, highlighting the degree of association between student behavior and HIV test results. A *p*-value < 0.05 was considered statistically significant, and confidence intervals were estimated at a 95% confidence level.

### Ethical Considerations

This study obtained ethical clearance from the Regional Ethics Committee for Human Health Research of the Centre (CRERSH-center) and from the Faculty of Medicine and Biomedical Sciences of the University of Yaoundé 1. Similarly, administrative authorizations were obtained from the chosen universities. The informed consent of the participant was systematically obtained prior data collection. All methods were performed in accordance with the relevant guidelines of the Helsinki declaration.

## Results

### Sociodemographic characteristics

All 1200-university students contacted for this study agreed to participate, resulting in a 100% response rate. However, some participants freely chose not to answer certain questions, hence the varying frequencies of responses depending on the question. According to the sociodemographic profile, one thousand one hundred and nineteen students were surveyed during this study, including 572 men and 628 women. Regarding the gender disparity, a slight overrepresentation of women (628) compared to men (572) is observed among the students surveyed in this study. The under 20-age group represented 49.0% of the sample, 21–24-year-olds 40%, and those 25 and over 11%. According to the level of education, 98% of students were enrolled in the first cycle, and 2% were attending the second cycle of university. The majority of students, 98%, lived outside the university campus, of whom 75% lived with family and 25% lived outside the family home with friends or in a student residence (Table 1).

**Table 1.**
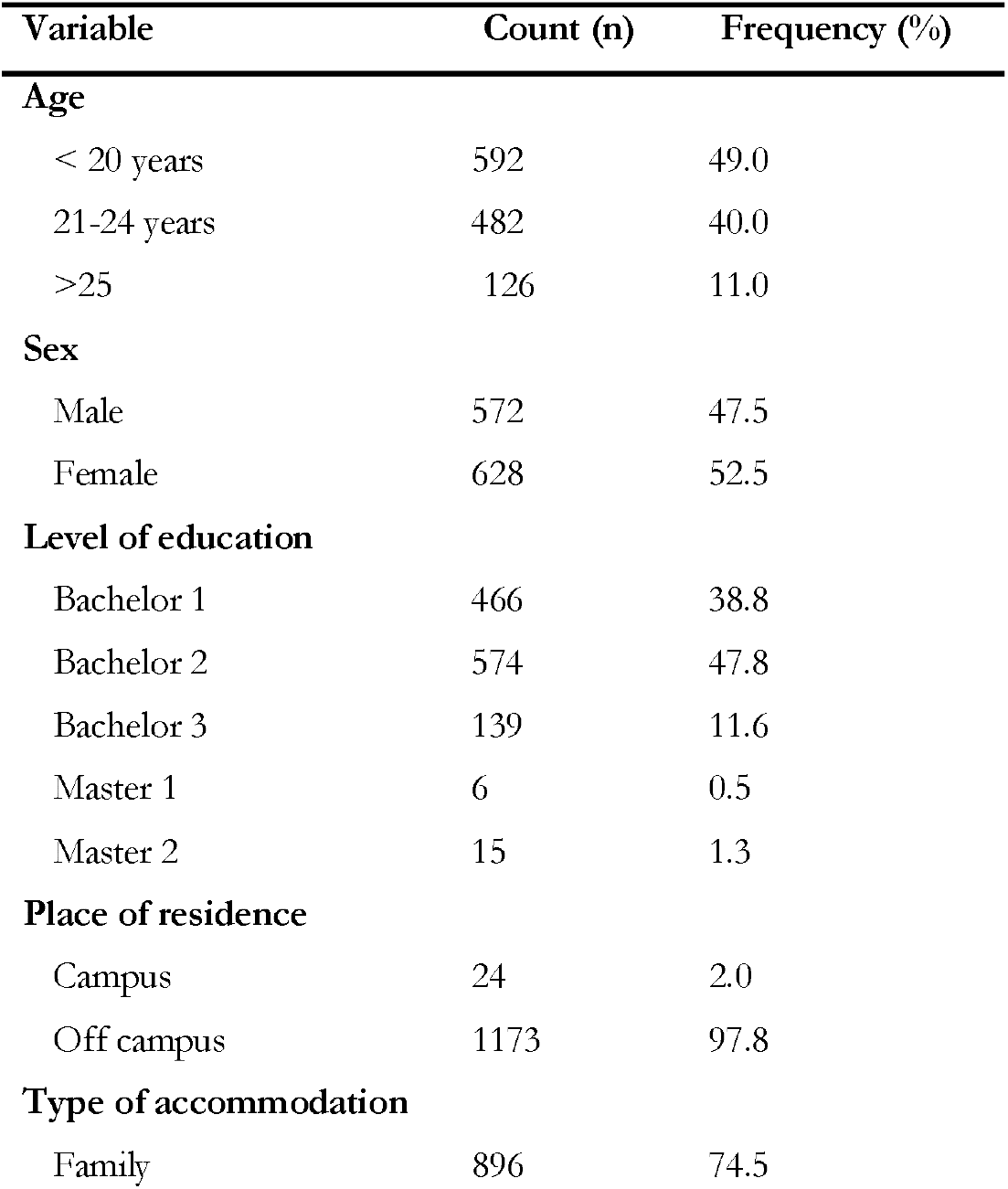
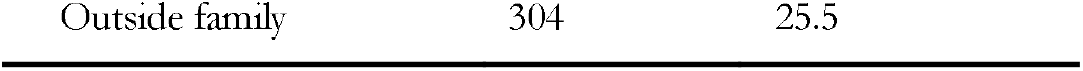
Sociodemographic profile of study participants, University of Yaoundé 1, Cameroon, 2024 (*n*=1200)

### Socioeconomic profile

Regarding socioeconomic characteristics, 92% of students had their parents as their main material/financial support. Among these students, 65% of this material support was regular, 48% found this support insufficient. Finally, 26% of students declared having a part-time job to be able to meet their academic needs. The socio-economic profile of the participants was heterogeneous (Table 2).

**Table 2.**
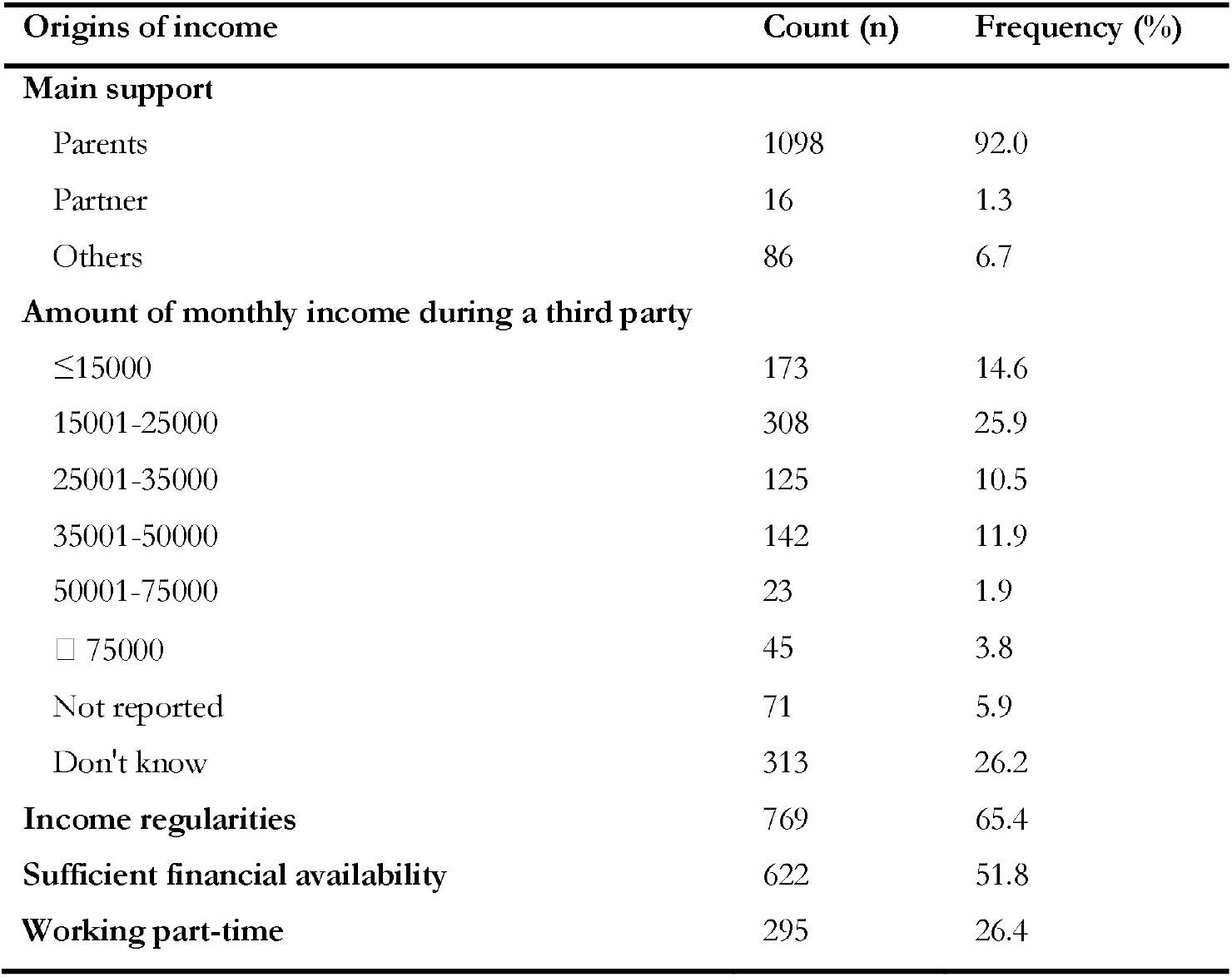
Socio-economic profile of study participants, University of Yaoundé 1, Cameroon, 2024.

### Sexual HIV risk behavior and factors related to HIV prevention

More than the majority of students, 69% of whom declared having had to consume alcohol during the last 12 months with 27% who consumed alcohol per month. A little more than half (57%) declared never having consumed a glass/bottle of alcohol per month, 18% a glass/bottle of alcohol once a month (Table 3).

**Table 3.** Alcohol consumption behavior for the last 12 months among students, University of Yaoundé 1, Cameroon, 2024 (*n*=1200)

| Variables | Count (n) | Frequency (%) |
| --- | --- | --- |
| **Alcohol consumption** |  |  |
| No | 251 | 30.1 |
| Yes | 574 | 68.7 |
| Not reported | 4 | 0.5 |
| Don't know | 6 | 07 |
| **Frequency of alcohol consumption** |  |  |
| Less than once a month | 116 | 15.1 |
| Once a month | 206 | 26.9 |
| 2-3 times a month | 196 | 25.6 |
| Once a week | 22 | 2.9 |
| 2-3 times a week | 9 | 1.2 |
| Every day or almost every day | 20 | 2.6 |
| Not reported | 91 | 11.9 |
| Don't know | 88 | 11.5 |
| **Frequency of drinking 6 glasses** |  |  |
| Never | 453 | 57.0 |
| Once a month | 140 | 17.6 |
| 2-3 times a month | 77 | 9.7 |
| Once a week | 14 | 1.8 |
| 2-3 times a week | 22 | 2.8 |
| Every day or almost every day | 12 | 1.5 |
| Not reported | 20 | 2.5 |
| Don't know | 57 | 7.2 |

### Comparison of sexual behaviors before and after implementation of interventions

The sample consisted of 528 men and 332 women in 2017 and 572 men and 628 women. Here, a significant increase in the number of women in 2024 compared to 2017 was observed, with a *p*-value □ 0.001. The low participation of men in this study can be explained by the fact that their use of available care activities and health services is generally limited, especially concerning sexual and reproductive health. Similarly, the most represented participants in this study belonged to the 18 to 24-year-old age group, with a significant increase in this age group in 2024 and a *p*-value ⍰ 0.001. Within our sample, compared to that of 2017, income-generating activities were heterogeneous with a *p*-value ⍰ 0.001(Table 4).

**Table 4.** Comparison of sociodemographic and socioeconomic profile of students before and after implementation of interventions, University of Yaoundé1, Cameroon, 2024 (*n*=1200)

| Characteristics | Pre-intervention<br>n=860 (%) | Post-intervention<br>n=1200 (%) | $p$-value |
| --- | --- | --- | --- |
| **Sex** |  |  |  |
| Male | 528(61.4) | 572(47.5) | $\leq 0.001$ |
| Female | 332(38.6) | 628(52.5) | $\leq 0.001$ |
| **Age groups** |  |  |  |
| 18-24 | 662(77.0) | 1179(98.3) | $\leq 0.001$ |
| 25-30 | 185(21.5) | 21(1.8) | $\leq 0.001$ |
| 31 + | 13(1.5) | 0.0(0.0) | 0.0 |
| **Place of residence** |  |  |  |
| On campus | 52(6.1) | 24(2.0) | $\leq 0.001$ |
| Off campus | 799(93.9) | 1173(97.8) | $\leq 0.001$ |
| **Family accommodation** |  |  |  |
| Yes | 465(62.3) | 896(74.5) | $\leq 0.001$ |
| No | 281(37.7) | 304(25.5) | $\leq 0.001$ |
| **Main support** |  |  |  |
| Parents | 749(89.6) | 1098(92.0) | $\leq 0.001$ |
| Partners | 32(3.8) | 16(1.3) | $\leq 0.001$ |
| State/stock market | 55(6.6) | 86(6.7) | $\leq 0.001$ |
| **Value of main monthly support** |  |  |  |
| <15,000 FCFA | 171(23.0) | 173(14.6) | $\leq 0.001$ |
| 15,001-25,000 FCFA | 117(15.7) | 308(25.9) | $\leq 0.001$ |
| 25,001-35,000 FCFA | 57(7.7) | 125(10.5) | $\leq 0.001$ |
| 35,001-50,000 FCFA | 47(6.3) | 142(11.9) | $\leq 0.001$ |
| 50,001-75,000 FCFA | 22(3.0) | 23(1.9) | $\leq 0.001$ |
| >75,000 FCFA | 22(3.0) | 45(3.8) | $\leq 0.001$ |
| Don't know (too variable) | 308(41.4) | 313(26.2) | $\leq 0.001$ |
| **Regularity of the financial market** |  |  |  |
| Yes | 437(61.6) | 769(65.4) | $\leq 0.001$ |
| No | 272(48.4) | 431(34.6) | $\leq 0.001$ |
| **Income earned part-time** |  |  |  |
| <15,000 FCFA | 64(35.6) | 78(23.7) | $\leq 0.001$ |
| 15,001-25,000 FCFA | 42(23.3) | 87(26.4) | $\leq 0.001$ |
| 25,001-35,000 FCFA | 25(13.9) | 24(7.3) | $\leq 0.001$ |
| 35,001-50,000 FCFA | 17(9.4) | 31(9.4) | $\leq 0.001$ |
| 50,001-75,000 FCFA | 6(3.3) | 13(4.0) | $\leq 0.001$ |
| Don't know (too variable) | 26(14.4) | 28(8.5) | $\leq 0.001$ |

The refinement of the analysis showed that certain subgroups have higher prevalences, notably young people who consume alcohol, although very few drink amounts capable of disorienting them regarding their school engagement (Table 5).

**Table 5.** Comparison between alcohol consumption and its consequences among students before and after the implementation of interventions, University of Yaoundé1, Cameroon, 2024 (*n*=1200)

| Characteristics | Pre-intervention<br>n=860 (%) | Post-intervention<br>n=1200 (%) | *p*-value |
| --- | --- | --- | --- |
| **How often do you drink alcohol?** |  |  |  |
| Once a month | 513(50.4) | 322(42.0) | □0.001 |
| 2-3 times a month | 164(16.1) | 196(25.6) | □0.001 |
| Once a week | 92(9.0) | 22(2.9) | □0.001 |
| 2-3 times a week | 26(2.6) | 9(1.2) | □0.001 |
| Every day or almost every day | 14(1.4) | 20(2.6) | □0.001 |
| Not reported | 41(4.0) | 91(11.9) | □0.001 |
| Don't know | 151(14.8) | 88(11.5) | □0.001 |
| **On one occasion, how many times can you drink six or more standard drinks?** |  |  |  |
| Never | 453(57.0) | 672(85.1) | □0.001 |
| Once a month | 140(17.6) | 3.3(26) | □0.001 |
| 2-3 times a month | 77(9.7) | 15(1.9) | □0.001 |
| Once a week | 14(1.8) | 3(0.4) | □0.001 |
| 2-3 times a week | 22(2.8) | 22(2.8) | □0.001 |
| Every day or almost every day | 12(1.5) | 7(0.9) | □0.001 |
| Not reported | 20(2.5) | 19(2.4) | □0.001 |
| Don't know | 57(7.2) | 48(6.1) | □0.001 |
| **In the past year, how many times, because you were drinking, were you unable to do what you were trying to do?** |  |  |  |
| Never | 533(52.4) | 673(84.8) | □0.001 |
| Once a month | 49(4.8) | 34(4.3) | □0.001 |
| 2-3 times a month | 14(1.4) | 15(1.9) | □0.001 |
| One a week | 16(1.6) | 11(1.4) | □0.001 |
| Every day or almost every day | 6(0.6) | 6(0.8) | □0.001 |
| Not reported | 19(1.9) | 13(1.6) | □0.001 |
| Don't know | 75(7.4) | 42(5.3) | □0.001 |
| **In the past year, how many times have you been unable to remember what happened the night before because you were drinking?** |  |  |  |
| Never | 879(86.4) | 672(85.1) | □0.001 |
| Once a month | 29(2.9) | 26(3.3) | □0.001 |
| 2-3 times a month | 16(1.6) | 15(1.9) | □0.001 |
| Once a week | 5(0.5) | 3(0.4) | □0.001 |
| Every day or almost every day | 2(0.2) | 7(0.9) | □0.001 |
| Not reported | 14(1.4) | 19(2.4) | □0.001 |
| Don't know | 45(4.4) | 48(6.1) | □0.001 |

Finally, we note that preventive practices among young people have seen notable progress between 2018 and 2023, thus highlighting the effectiveness of prevention strategies, in this case awareness raising and accessibility to condoms. From a programmatic point of view, this result is generally positive and proves that the results of prevention in the fight against AIDS have progressed on campuses, according to the analysis of the Effects of the interventions carried out. On the other hand, between 2020-2023, the proportion of young people who did not use condoms remained stable. As young people residing in the family home and the most educated have contributed more to maintaining risky behaviors, it appears necessary to redefine the objectives and reformulate prevention programs with an emphasis on the use of condoms (Table 6).

**Table 6.** Comparison between sexual behaviors regarding STIs among students before and after implementation of interventions, University of Yaoundé1, Cameroon, 2024 (*n*=1200)

| Indicator | Pre-intervention<br>n=860 (%) | Post-intervention<br>n=1200 (%) | *p*-value |
| --- | --- | --- | --- |
| **People reporting an experienced STI symptom** |  |  |  |
| Feeling of pain or burning when you urinate | 358(35.2) | 75(19.3) | □0.001 |
| Unusual vaginal discharge or white spotting or irregular bleeding from the vagina | 358(35.2) | 55(14.1) | □0.001 |
| Genital warts | 43(4.2) | 10(2.6) | □0.001 |
| Anal warts | 43(4.2) | 3(0.8) | □0.001 |
| Abnormal mass or swelling around the genital tract | 98(9.7) | 16(4.1) | □0.001 |
| Tingling | 19(1.9) | 49(12.6) | 0.014 |
| No | 473(46.5) | 181(46.5) | □0.001 |
| **Management and treatment of an STI** |  |  |  |
| Yes, under the care of a doctor or other healthcare personnel | 421(41.1) | 122(41.2) | □0.001 |
| Yes, taken care of by a traditional practitioner | 34(3.4) | 11(3.7) | □0.001 |
| No, they were not supported | 270(24.6) | 33(11.1) | □0.001 |
| No, I handled them myself | 147(14.5) | 24(8.1) | □0.001 |
| They were milked by someone else | 29(2.9) | 0(0) | 0.0 |
| Refusal | 64(6.3) | 55(18.6) | □0.001 |
| Don't know | 73(7.2) | 51(17.2) | □0.001 |
| **Suggested use of condoms** |  |  |  |
| No | 78(7.7) | 110(14.5) | □0.001 |
| Yes | 931(91.5) | 651(85.5) | □0.001 |
| **Has already used a condom** |  |  |  |
| No | 76(7.5) | 165(20.1) | □0.001 |
| Yes | 931(91.5) | 656(79.9) | □0.001 |
| **Condom accessibility** |  |  |  |
| Very difficult | 134(13.2) | 67(5.6) | □0.001 |
| Quite difficult | 67(6.6) | 41(3.4) | □0.001 |
| Neither difficult nor easy | 220(21.6) | 228(19.0) | □0.001 |
| Quite easy | 200(19.7) | 182(15.2) | □0.001 |
| Very easy | 395(38.8) | 100(8.3) | □0.001 |
| **Difficulty obtaining condoms** |  |  |  |
| Not easy to find condoms | 207(20.4) | 55(15.3) | □0.001 |
| Worried that other people would go see him get them | 114(11.2) | 50(13.9) | □0.001 |
| Too expensive | 99(9.7) | 18(5.0) | □0.001 |
| Other | 32(3.1) | 31(8.6) | □0.001 |
| Not reported | 176(17.3) | 77(21.4) | □0.001 |
| Don't know | 385(38.3) | 129(35.8) | □0.001 |
| **Source of supply of condoms** |  |  |  |
| Public place | 127(12.5) | 15(4.7) | □0.001 |
| Shops | 699(68.7) | 226(70.2) | □0.001 |
| Health facilities | 322(31.7) | 59(18.3) | □0.001 |
| others | 20(2) | 22(6.8) | □0.001 |

**Table 7.** Comparison between alcohol consumption and its consequences among students after the implementation of interventions, University of Yaoundé1, Cameroon, 2024 (*n*=1200)

| Indicator | Post-intervention<br>n (%) | Frequency<br>(%) |
| --- | --- | --- |
| **How many standard drinks do you have on a typical drinking day?** |  |  |
| 1-2 | 0 | 0 |
| 3-4 | 673 | 84.8 |
| 5-6 | 34 | 4.3 |
| 7-9 | 15 | 1.9 |
| 10+ | 11 | 1.4 |
| Not reported | 6 | 0.8 |
| Don't know | 55 | 6.9 |
| **In the past year, how many times have you found yourself unable to stop drinking after you started?** |  |  |
| Never | 672 | 85.1 |
| Once a month | 26 | 3.3 |
| 2-3 times a month | 15 | 1.9 |
| Once a week | 3 | 0.4 |
| 2-3 times a week | 22 | 2.8 |
| Every day or almost every day | 7 | 0.9 |
| Not reported | 19 | 2.4 |
| Don't know | 48 | 6.1 |
| **In the past year, how many times, after a period of heavy drinking, have you had to drink alcohol first thing in the morning to get back in shape?** |  |  |
| Never | 673 | 84.8 |
| Once a month | 34 | 4.3 |
| 2-3 times month | 15 | 1.9 |
| Once a week | 11 | 1.4 |
| Every day or almost every day | 6 | 0.8 |
| Not reported | 13 | 1.6 |
| Don't know | 42 | 5.3 |
| **In the past year, how often have you felt guilty or regretful after drinking?** |  |  |
| Never | 673 | 84.8 |
| Once a month | 34 | 4.3 |
| 2-3 times a month | 15 | 1.9 |
| Once a week | 11 | 1.4 |
| Every day or every day | 6 | 0.8 |
| Not reported | 13 | 1.6 |
| Don't know | 42 | 5.3 |
| **Have you hurt yourself or someone else because you were drinking?** |  |  |
| No | 1197 | 100 |
| Yes, but not in the last year | 0 | 0 |
| Yes, in the last year | 0 | 0 |
| **In the past year, how often have you felt guilty or regretful after drinking?** |  |  |
| No | 1197 | 100 |
| Yes, but not in the last year | 0 | 0 |
| Yes, in the last year | 0 | 0 |

### Link between student behavior and HIV test results

Regarding the link between student behavior and HIV test results, it appeared that we cannot precisely identify the prevalence that is truly considered a factor that can explain a positive HIV test result within the month. Because the prevalence represented a rate of 25%.

## Discussion

### Participant Profile

Of the 1197 students surveyed, 572 were men and 628 were women, giving a male-to-female ratio of 0.91. The slight overrepresentation of women in our sample reflects the demographics of the Cameroonian population and their more frequent use of health services, particularly in sexual and reproductive health [33]. The majority of participants (100%) were under 35 years old, which corresponds to the [15-49 years] age group predominant in HIV testing and treatment activities. These results are similar to those of previous studies conducted in Yaoundé [2], but differ from those conducted in Rwanda, where older students had better knowledge of HIV/AIDS. These studies suggest a positive correlation between age and HIV/AIDS knowledge.

Regarding the place of residence, 97.8% of students lived off-campus, with 74.5% living with family and 25.5% living outside the family home. Living with parents promotes parental control and reduces the likelihood of risky sexual behavior [34]. Studies have shown that living in a family, whether extended or nuclear, is associated with a later age of first sexual intercourse [34]. Parent-child dialogue on health is also crucial, as it allows young people to be better informed about prevention [33]. However, other studies have found different results regarding on-campus residence and HIV testing [35]. It is therefore important to encourage parents to discuss sexuality with their children [36].

Income-generating activities were heterogeneous in our sample. The majority of students (92.0%) were financially supported by their parents, while 1.3% were supported by their partners and 6.7% by the state or a scholarship. These results are consistent with the 2017 IBBS surveys in Cameroon, which did not find a significant link between economic conditions and first sexual intercourse [37]. However, adolescents in difficult economic situations are more likely to have multiple sexual partners [37]. The World Bank (2001) highlighted the correlation between poverty, inequality and HIV infection rates, as well as the economic pressure that can push young girls to accept sexual intercourse in exchange for support [38]. Rwenge’s study (reference) also revealed a strong correlation between economic factors and risky sexual behaviors.

### Alcohol Consumption and Risky Behaviors

Alcohol consumption is a significant risk factor for risky sexual behaviors. Just over half of the students consume alcohol once a month, although only 8.5% consume amounts that could impair their judgment. These results are similar to those of Billong et al. (2017), which showed that alcohol and drug use is associated with risky sexual behaviors [2]. Alcohol impairs judgment, which can lead to unprotected sex and an increased risk of HIV infection. A narrative review of 86 studies by Blignaut RJ (reference) confirmed that people with a history of alcohol consumption and those whose partners regularly consume alcohol are more at risk of HIV infection [39]. Similarly, Kagou et al.’s study revealed that alcohol consumption by young girls or their partners reduces condom use during sexual intercourse [26].

Participation in concerts is also associated with risky behaviors, likely due to excessive alcohol and drug consumption [40]. However, in our sample, we did not find a statistically significant association between alcohol consumption and positive HIV test results. Other studies have shown that students do not consistently use condoms, especially in stable relationships based on trust [41]. The high prevalence of HIV may be related to risky behaviors such as sexual intercourse with sex workers and inconsistent condom use. The presence of tourists, nightclubs, and guest houses can also encourage transactional sex [42]. Our results confirm the association between early sexual intercourse and an increased risk of HIV infection [43]. Psychoactive substance use can lead to loss of inhibition and risky sexual behaviors [42].

### STIs and Prevention

STIs are a major health problem in Africa, as they increase the risk of HIV transmission. Most cases are neither diagnosed nor treated, and studies often focus on specific subpopulations [26]. In our study, more than half of the students reported STI symptoms in the past 12 months, which is higher than the results of Billong et al. (25.1%) [2]. This could be due to limited access to STI testing and treatment services.

In Africa, HIV/STI transmission is mainly sexual, and young people aged 15 to 29 are the most affected. Prevention programs therefore target adolescents and young people, considered a risk group. In our sample, consistent condom use was common among 21-24 year olds (80%), which is consistent with the results of Billong et al. (92%) [2]. However, condom use at first sexual intercourse remains low [33].

African social norms mean that the suggestion to use a condom often falls to men [26]. In our study, 86% of students have already suggested condom use, which is similar to the results of the 2017 IBBS [2].

Most students in our study used condoms during sexual intercourse, which is consistent with other research [40]. Drug and alcohol abuse is associated with risky sexual behaviors, as it can impair judgment and lead to unprotected sex [40]. However, a study showed that among students over 24, ignorance of serological status is a major problem [3]. The number of students reporting STI symptoms is inconsistent with high condom use, suggesting problems with correct use [2]. HIV testing was less common among men than among women, which is consistent with other studies [40]. It is therefore important to strengthen HIV testing awareness campaigns [44].

### Study limitations

Like any study, ours has limitations. The cross-sectional design of our study does not allow us to establish causal relationships. In addition, the slowness of administrative procedures hindered the progress of our work. The reluctance of some students to participate also limited our sample. Finally, data collection in a single region may limit the generalizability of our results.

## Conclusions

It appears necessary to redefine the objectives and reformulate prevention programs by strengthening the implemented strategies that have proven effective; however, this should be done with an emphasis on condom use.

## Data Availability

All data produced in the present work are contained in the manuscript and its supplemental materials

## Declaration

## Author’s contribution

Drafting of the protocol, data collection, analysis and interpretation: MA, EOG, HY, LE, TM and SCB: Drafting of original manuscript: MA, FZLC, LE, TM and SCB; Critical revision of the manuscript: FZLC, LE, TM and SCB; Conception, design, supervision of implementation, editing and final validation of the manuscript: MA, LE, TM and SCB. All authors approved the final version of the article.

## Ethical Approval Statement

Institutional Review Board (IRB) of the Faculty of Medicine and Biomedical Sciences of Yaoundé approved the protocol and the ethical clearance issued. Informed consent was obtained from participants prior to inclusion in the study.

## Consent for publication

Not applicable.

## Availability of data and materials

All data generated or analyzed during this study are included in this published article and supplementary materials.

## Acknowledgements

We would like to thank all the students who participated in this study and the managers of the establishments concerned who authorized this study to be carried out in their establishments.

