## Additional Files for "Effects of HIV prevention interventions on the behaviors of students at the university campuses of Yaoundé (Cameroon)"

### **Identify HIV interventions implemented between 2017-2023**

**Number of PLHIV in Cameroon**

In Cameroon, the number of PLHIV estimated in 2023 was 474,764 (HIV Estimates and Projections Report 2023), including 26,182 (5.5%) children under 15 and 317,108 (66.8%) women.

Numbers of PLVIH

| Years | PLHIV (total) |
| --- | --- |
| 2019 | 491894 |
| 2020 | 488640 |
| 2021 | 484869 |
| 2022 | 480228 |
| 2023 | 474764 |

**Fig.1** Estimated number of PLHIV total in Cameroon from 2018-2023

*Source: HIV Estimates and Projections Report 2023; PLVIH: People living with HIV*

**Fig. 2-Estimated** total numbers of children less than 15 years old in Cameroon from 2018-2023

numbers of children <16 yers

| Years | Children < 15 years old |
| --- | --- |
| 2019 | 35388 |
| 2020 | 33475 |
| 2021 | 31280 |
| 2022 | 29168 |
| 2023 | 26182 |

*Source: HIV Estimates and Projections Report 2023*

Numbers of adolescents 15-19 years

| Years | Adolescents (15-19 years old) |
| --- | --- |
| 2019 | 19012 |
| 2020 | 18351 |
| 2021 | 17632 |
| 2022 | 16719 |
| 2023 | 15751 |

**Fig.3** Estimated total numbers of Adolescents 15-19 years old in Cameroon from 2018-2023

*Source: HIV Estimates and Projections Report 2023*

young People 20-24 years

| Years | Young people (20-24 years old) |
| --- | --- |
| 2019 | 32732 |
| 2020 | 31152 |
| 2021 | 29560 |
| 2022 | 27796 |
| 2023 | 26182 |

**Fig.4** Estimated total numbers of young people 20-24 old in Cameroon from 2018-2023.

*Source: HIV Estimates and Projections Report 2023*

Adults 15-45 ans

| Years | Adults (15-49 years) |
| --- | --- |
| 2019 | 382192 |
| 2020 | 376163 |
| 2021 | 368826 |
| 2022 | 359988 |
| 2023 | 350477 |

**Fig.5:** Estimated total numbers of Adults 15-49 old in Cameroon from 2018-2023

*Source: HIV Estimates and Projections Report 2023*

Women

| Years | Women |
| --- | --- |
| 2019 | 323096 |
| 2020 | 322598 |
| 2021 | 321509 |
| 2022 | 319583 |
| 2023 | 317108 |

**Fig.6:** Estimated total numbers of Women in Cameroon from 2018-2023

*Source: HIV Estimates and Projections Report 2023*

**Number of new HIV infections**

Reduction in new infections: Recent estimates show a drop in new infections in the general population of 55% between 2019 and 2023 (from 16,311 in 2019 to 7,297 in 2023).

Among children under 15, this drop is almost 35.2% (from 4,831 in 2019 and 1,702 in 2022). Women accounted for almost 2/3 (65%) of new infections.

new infection

| Years | New infections (total) |
| --- | --- |
| 2019 | 16311 |
| 2020 | 14074 |
| 2021 | 12221 |
| 2022 | 9898 |
| 2023 | 7297 |

**Fig.7:** Total numbers of new HIV infections in Cameroon from 2018-2023

*Source: HIV Estimates and Projections Report 2023*

Childrens <15 old

| Years | childrens < 15 old |
| --- | --- |
| 2019 | 4831 |
| 2020 | 4411 |
| 2021 | 3793 |
| 2022 | 3414 |
| 2023 | 1702 |

**Fig.8:** Total numbers of new HIV infections of children’s in Cameroon from 2018-2023

*Source: HIV Estimates and Projections Report 2023*

Adolescent

| Years | Adolescents (15-19 old) |
| --- | --- |
| 2019 | 2125 |
| 2020 | 1797 |
| 2021 | 1577 |
| 2022 | 1218 |
| 2023 | 1052 |

**Fig.9:** Total numbers of new HIV infections of adolescent in Cameroon from 2018-2023

*Source: HIV Estimates and Projections Report 2023*

Young peaple

| Years | Young people (20-24 old) |
| --- | --- |
| 2019 | 2426 |
| 2020 | 2031 |
| 2021 | 1766 |
| 2022 | 1359 |
| 2023 | 1176 |

**Fig.10:** Total numbers of new HIV infections of young people in Cameroon from 2018-2023

*Source: HIV Estimates and Projections Report 2023*

Adults

| Years | Adults (15-49 old ) |
| --- | --- |
| 2019 | 10708 |
| 2020 | 9015 |
| 2021 | 7862 |
| 2022 | 6047 |
| 2023 | 5217 |

**Fig.11:** Total numbers of new HIV infections of Adults in Cameroon from 2018-2023

*Source: HIV Estimates and Projections Report 2023*

Women

| Years | Women |
| --- | --- |
| 2019 | 10394 |
| 2020 | 8949 |
| 2021 | 7799 |
| 2022 | 6256 |
| 2023 | 4792 |

**Fig.12:** Total numbers of new HIV infections of Women in Cameroon from 2018-2023

*Source: HIV Estimates and Projections Report 2023*

**Measures to prevent HIV infection**

Distribution of condoms, lubricating gels and prevention among KPs:

During the year 2023, approximately 28,180,438 male condoms, 1,005,380 female condoms, and 5,615,620 lubricants were distributed [in Cameroon]. The populations most at risk of HIV infection were the primary beneficiaries.

The distribution of these products mainly occurred during HIV awareness activities and campaigns. Pre-Exposure Prophylaxis (PrEP) is a complementary biomedical HIV prevention strategy whose principle is to give ARVs to uninfected (HIV-negative) individuals before exposure to HIV, in the context of high-risk sexual relationship(s).

During the year 2023 [in Cameroon], PrEP dispensing activities were carried out by several partners, including CARE and CHP. The following results were obtained along the continuum of prevention care offered to key populations (MSM and SW).

Female condoms distributed

| Years | Female condoms |
| --- | --- |
| 2019 | 3401584 |
| 2020 | 2918908 |
| 2021 | 2660738 |
| 2022 | 2846635 |
| 2023 | 1005380 |

**Fig.13:** Numbers of female condoms distributed in Cameroon from 2018-2023

*Source: CARE and CHP 2023 activity report and CNLS 2022 annual report*

Male condoms

| Years | Male condoms |
| --- | --- |
| 2019 | 34978318 |
| 2020 | 41057011 |
| 2021 | 33111351 |
| 2022 | 29122272 |
| 2023 | 28180438 |

**Fig.14:** Numbers of Male condoms distributed in Cameroon from 2018-2023

*Source: CARE and CHP 2023 activity report and CNLS 2022 annual report*

Lubricating gels

| Years | Lubricating Gels |
| --- | --- |
| 2019 | 4791302 |
| 2020 | 912714 |
| 2021 | 14059136 |
| 2022 | 12347440 |
| 2023 | 0 |

**Fig.15:** Numbers of lubricating gels condoms distributed in Cameroon from 2018-2023

*Source: CARE and CHP 2023 activity report and CNLS 2022 annual report*

Prep among MSM

| Years | Prep among MSM |
| --- | --- |
| 2020 | 1064 |
| 2021 | 1376 |
| 2022 | 1853 |
| 2023 | 2020 |

**Fig.16:** Numbers of prep among MSM distributed in Cameroon from 2018-2023

*Source: CARE and CHP 2023 activity report and CNLS 2022 annual report*

Prep for TS

| Years | Prep for TS |
| --- | --- |
| 2020 | 688 |
| 2021 | 1082 |
| 2022 | 2984 |
| 2023 | 3789 |

**Fig.17:** Numbers of prep for TS distributed in Cameroon from 2018-2023

*Source: CARE and CHP 2023 activity report and CNLS 2022 annual report*

**Screening.**

It remained in constant evolution throughout the period, increasing from 59.5% in 2017 to 98.1% in 2023. This performance is the result of case identification strategies through active community outreach, index testing, self-testing, and screening at all entry points of health facilities for both adult and pediatric populations. Consequently, the achievement of the first 95 is effective in the Cameroonian context, highlighting the need to sustain these good practices and intensify interventions among children and adolescents, for whom performance remains suboptimal.

People tested

| Years | Number of people tested |
| --- | --- |
| 2019 | 2941083 |
| 2020 | 2984346 |
| 2021 | 3479989 |
| 2022 | 2948355 |
| 2023 | 2638708 |

**Fig.18:** Numbers of people tested distributed in Cameroon from 2018-2023

*Source: FOSA monthly activity reports in 2023 and CNLS 2022 annual report*

People tested HIV+

| Years | Number of people tested HIV+ |
| --- | --- |
| 2019 | 83243 |
| 2020 | 99273 |
| 2021 | 92829 |
| 2022 | 70206 |
| 2023 | 54916 |

**Fig.19:** Numbers of people tested HIV+ in Cameroon from 2018-2023

*Source: FOSA monthly activity reports in 2023 and CNLS 2022 annual report*

HIV rate

| Years | HIV rate |
| --- | --- |
| 2019 | 2.8% |
| 2020 | 3.3% |
| 2021 | 2.7% |
| 2022 | 2.4% |
| 2023 | 2.1% |

**Fig.20:** Numbers of HIV rate in Cameroon from 2018-2023

*Source: FOSA monthly activity reports in 2023 and CNLS 2022 annual report*

**Support for PLHIV**

Link to ART

Overall, a stagnation in the rate of linkage to treatment has been observed across all age groups over the last 3 years. Nevertheless, the lowest rates remain observed among young people and adolescents (10-14 years and 15-19 years). The reasons that would explain this poor outcome among children include the difficulty in having an adult (parent, guardian) for their follow-up. Furthermore, there is the low geographical coverage of pediatric care sites and the insufficient number of providers trained in the disclosure of serological status and the management of children and adolescents. In 2023, linkage to ART among key populations ranged from 72.6% among SWs to 88.6% among MSM (figure below). Although progress has been increasing over the past few years, a slight decrease was observed in all subgroups in 2023. Linkage to ART among vulnerable populations, although showing increasing trends over time, remains below the national average for all categories, ranging from 69.7% among CSW to 79.8% among AGYW.

Sex Workers

| Years | Sex workers |
| --- | --- |
| 2019 | 80.1% |
| 2020 | 64.1% |
| 2021 | 75.1% |
| 2022 | 78.9% |
| 2023 | 72.6% |

**Fig.21:** Linkage to ART among key and vulnerable populations in Cameroon from 2018-2023: Sex Workers

*Source: CARE and CHP activity report in 2023 and CNLS 2022 annual report*

HSH

| Years | HSH |
| --- | --- |
| 2019 | 76.7% |
| 2020 | 42.2% |
| 2021 | 80.7% |
| 2022 | 95.1% |
| 2023 | 88.6% |

**Fig.22:** Linkage to ART among key and vulnerable populations in Cameroon from 2018-2023: HSH

*Source: CARE and CHP activity report in 2023 and CNLS 2022 annual report*

UDI

| Years | UDI |
| --- | --- |
| 2019 | 0% |
| 2020 | 68.9% |
| 2021 | 75.9% |
| 2022 | 82.2% |
| 2023 | 77.6% |

**Fig.23:** Linkage to ART among key and vulnerable populations in Cameroon from 2018-2023: UDI

*Source: CARE and CHP activity report in 2023 and CNLS 2022 annual report*

TG

| Years | TG |
| --- | --- |
| 2019 | 64.3% |
| 2020 | 54% |
| 2021 | 85.1% |
| 2022 | 85.6% |
| 2023 | 81% |

**Fig.24:** Linkage to ART among key and vulnerable populations in Cameroon from 2018-2023: TG

*Source: CARE and CHP activity report in 2023 and CNLS 2022 annual report*

Sex Workers customers

| Years | Sex workers customers |
| --- | --- |
| 2019 | 61.2% |
| 2020 | 71.3% |
| 2021 | 60.6% |
| 2022 | 65.1% |
| 2023 | 69.7% |

**Fig.25:** Linkage to ART among key and vulnerable populations in Cameroon from 2018-2023: Sex Workers Customers

*Source: CARE and CHP activity report in 2023 and CNLS 2022 annual report*

JFNS

| Years | JFNS |
| --- | --- |
| 2019 | 66.8% |
| 2020 | 67.9% |
| 2021 | 66.3% |
| 2022 | 67.3% |
| 2023 | 74.3% |

**Fig.26:** Linkage to ART among key and vulnerable populations in Cameroon from 2018-2023: JFNS

*Source: CARE and CHP activity report in 2023 and CNLS 2022 annual report*

JGNS

| Years | JGNS |
| --- | --- |
| 2019 | 71.7% |
| 2020 | 67.6% |
| 2021 | 70.1% |
| 2022 | 69.1% |
| 2023 | 79.8% |

**Fig.27:** Linkage to ART among key and vulnerable populations in Cameroon from 2018-2023: JGNS

*Source: CARE and CHP activity report in 2023 and CNLS 2022 annual report*

**Children under 15 years old, adolescents, and youth on ART**

Overall, there is an upward trend in the active patient caseload for both children under 15 years and adults 15 years and older, although the rate differs between the two groups.

Regarding the adult caseload, it experienced an average annual growth rate of 9.33% between 2020 and 2023, increasing from 339,599 to 437,470. This increase in the active caseload is likely due to the reduction in lost to follow-up (LTFU) cases, thanks to various tracking strategies for LTFU patients and those absent from treatment, on the one hand, and on the other hand, the implementation of differentiated ART delivery strategies in the community.

As for the pediatric caseload, it experienced a growth rate of 2.7% from 2020 to 2021, increasing from 11,219 to 11,531. Between 2022 and 2023, a slight decrease was noted, which could be explained by the migration of 14-year-old children to the adult caseload and, as observed above, the decrease in the number of new enrollments.

Children under 15 on ART

| Years | Children under 15 years old |
| --- | --- |
| 2019 | 25.7% |
| 2020 | 32.5% |
| 2021 | 34.9% |
| 2022 | 39.5% |
| 2023 | 40.9% |

**Fig.28:** Children under 15 on ART in Cameroon from 2018-2023

*Source: Monthly activity report of the FOSA in 2023 and annual report of the CNLS 2022*

Adolescents on ART

| Years | Adolescents aged 10 -19 |
| --- | --- |
| 2019 | 32.2% |
| 2020 | 35.2% |
| 2021 | 39% |
| 2022 | 45.6% |
| 2023 | 46.5% |

**Fig.29:** Adolescents aged 10-19 years on ART in Cameroon from 2018-2023

*Source: Monthly activity report of the FOSA in 2023 and annual report of the CNLS 2022*

Young Peaple on ART

| Years | Young people aged 20 -24 |
| --- | --- |
| 2019 | 48.3% |
| 2020 | 60.7% |
| 2021 | 66.4% |
| 2022 | 81.5% |
| 2023 | 83% |

**Fig 30:** Young People aged 20-24 years on ART in Cameroon from 2018-2023

*Source: Monthly activity report of the FOSA in 2023 and annual report of the CNLS 2022*

**Follow-up of PLHIV on ART**

Initiation of ART for PLHIV identified: an overall improvement of 17.4% in ART coverage is observed. This performance was facilitated by the effective implementation of the "Test and Treat" strategy, task shifting, and the decentralization of treatment provision.

Load Testing Coverage: Despite a slight increase, access to VL remains below the minimum recommended threshold throughout the 2017-2023 period, with a drastic decrease in 2023 (45.3%). Although the intermittent availability of reagents is the main reason justifying this poor performance, it is important to highlight the challenges in the supply chain of inputs between the central level and reference laboratories, the need for optimization of the mapping for VL coverage, and the integration of all existing platforms for conducting VL tests within the framework of health system strengthening.

Viral Suppression: Despite the progress observed between 2017 and 2019 (from 50.1% to 88.0%), viral load suppression has stagnated around 85%. This is a result of challenges in identifying non-suppressed patients (for adherence reinforcement) or those experiencing treatment failure (for timely switching to a new treatment regimen). Thus, a consistent supply of VL tests, coupled with a continuous transition to dolutegravir-based treatment regimens, would enable the achievement of the third 95.

PLHIV identified

| Years | PLHIV identified |
| --- | --- |
| 2019 | 312214 |
| 2020 | 413188 |
| 2021 | 469783 |
| 2022 | 460088 |
| 2023 | 481147 |

**Fig.31:** Monitoring of PLHIV on ART in Cameroon from 2018-2023: PLHIV identified

*Source: database of reference laboratories in 2023 and CNLS 2022 annual report*

percnetage awareness of status

| Years | % awareness of status |
| --- | --- |
| 2019 | 83.8% |
| 2020 | 83.2% |
| 2021 | 94.1% |
| 2022 | 95.8% |
| 2023 | 98.1% |

**Fig. 32:** Monitoring of PLHIV on ART in Cameroon from 2018-2023: percentage awareness of status

*Source: database of reference laboratories in 2023 and CNLS 2022 annual report*

percentage link to ART

| Years | % link to ART |
| --- | --- |
| 2019 | 73.7% |
| 2020 | 84.9% |
| 2021 | 82.7% |
| 2022 | 93.3% |
| 2023 | 96.2% |

**Fig. 33:** Monitoring of PLHIV on ART in Cameroon from 2018-2023: percentage link to ART

*Source: database of reference laboratories in 2023 and CNLS 2022 annual report*

CV produced

| Years | Number of CVs produced |
| --- | --- |
| 2019 | 107506 |
| 2020 | 203905 |
| 2021 | 241613 |
| 2022 | 265019 |
| 2023 | 203134 |

**Fig. 34:** Monitoring of PLHIV on ART in Cameroon from 2018-2023: Number of CVs produced

*Source: database of reference laboratories in 2023 and CNLS 2022 annual report*

percentage CV achievement

| Years | % CV achievement |
| --- | --- |
| 2019 | 34.4% |
| 2020 | 58.1% |
| 2021 | 62.2% |
| 2022 | 62.4% |
| 2023 | 45.3% |

**Fig. 35:** Monitoring of PLHIV on ART in Cameroon from 2018-2023: Percentage CV achievement

*Source: database of reference laboratories in 2023 and CNLS 2022 annual report*

:

CVs deleted

| Years | CVs deleted |
| --- | --- |
| 2019 | 94617 |
| 2020 | 172212 |
| 2021 | 227429 |
| 2022 | 236493 |
| 2023 | 174593 |

**Fig. 36:** Monitoring of PLHIV on ART in Cameroon from 2018-2023: CVs deleted

*Source: database of reference laboratories in 2023 and CNLS 2022 annual report*

percnetage viral suppression

| Years | % viral suppression |
| --- | --- |
| 2019 | 88% |
| 2020 | 84.5% |
| 2021 | 94.1% |
| 2022 | 89.2% |
| 2023 | 85.9% |

**Fig. 37:** Monitoring of PLHIV on ART in Cameroon from 2018-2023: Percentage viral suppression

*Source: database of reference laboratories in 2023 and CNLS 2022 annual report*

retention at 12 months

| Years | Retention at 12 months |
| --- | --- |
| 2019 | 73.7% |
| 2020 | 75.7% |
| 2022 | 85% |
| 2023 | 87% |

**Fig. 38:** Monitoring of PLHIV on ART in Cameroon from 2018-2023: Retention at 12 months

*Source: database of reference laboratories in 2023 and CNLS 2022 annual report*

Status awareness 98.1%

ARV treatment 93.3%

Viral Removal: 85.9%
